# Multimodal artificial intelligence using entire electronic health record and complete pathogen genome data for patient outcome prediction from life-threatening infection: the SuperbugAI Platform

**DOI:** 10.64898/2026.08.04.26359536

**Authors:** Sonika Tyagi, Yashpal Ramakrishnaiah, Jane Hawkey, Jess Wisniewski, Luke Blakeway, Theo Christian, Vito Sikiric, William Librata, Jiangning Song, Geoffrey I. Webb, Aadith Ashok, Christopher Bain, Nenad Macesic, Anton Y. Peleg

## Abstract

Artificial intelligence (AI) has the potential to transform healthcare, with advanced multimodal approaches showing great promise in leveraging diverse health-related data. Here, we applied multimodal AI to entire electronic health record (EHR) and complete pathogen genome data to predict patient outcomes from life-threatening infection. An automated, scalable pipeline was developed for EHR data preprocessing, quality control, and standardisation. A deep learning fusion model was trained to predict in-hospital mortality, need for ICU admission, prolonged length of stay and 30-day unplanned readmission. We then developed a novel genomic large language model (gLLM) architecture to incorporate bacterial genomic features into the multimodal fusion model. The cohort comprised 2,656 bloodstream infection hospitalisations involving 2,535 patients. Deep learning fusion models using entire structured and unstructured EHR data outperformed traditional APACHE II score mortality prediction (AUROC [95% confidence intervals] 0.93 [0.92-0.94] versus 0.77 [0.77–0.78]). The model also showed strong performance for predicting the need for ICU admission (AUROC 0.978 [0.966 - 0.986]), prolonged hospital length of stay (AUROC 0.803 [0.790 - 0.812]) and unplanned readmission (AUROC 0.696 [0.690 - 0.701]). As proof of principle, incorporating entire microbial genomic features from the causative pathogen further enhanced prediction and enabled identification of key bacterial virulence pathways relevant for human disease. Multimodal AI integrating harmonised EHR and genomic data can accurately identify hospitalised patients at risk of poor outcomes. These approaches are scalable to other subspecialities of medicine.

## Introduction

The advent of electronic health records (EHR) has led to a massive expansion of diverse and complex healthcare data.^1,2^ The technology of artificial intelligence (AI) holds promise to leverage this explosion in health-related data and transform the practice of medicine.^3–5^ AI systems have the capacity to process, analyse and output results from billions of data points, and the sophistication of these systems to handle more diverse and complex data inputs is advancing rapidly.^6–8^ However, challenges exist for effective translation and implementation of AI tools into real-world clinical settings,^9,10^ including the ability to ingest data from highly diverse and disorganised hospital data warehouses and the processing, standardisation and input of alternate data sources (e.g. genomics) for clinically relevant patient outcome prediction.

Unlike many other subspecialities in medicine, human diseases caused by infections are determined by more than just anomalies of the human host. The properties of the microbial pathogen, and their interactions with the unique characteristics of the human host, plays a major role in the manifestations, severity and outcomes of disease.^11^ This complex interplay highlights the potential power of AI and precision medicine to improve the care of patients with infectious diseases. The pathogen genome adds an exciting new dimension that may augment prediction of clinical outcomes and support optimised care and personalised treatment. Features within the microbial genome may encode for factors that drive disease severity and patient survivability, pathogen transmission risk and outbreak potential, and degree of antimicrobial resistance and treatment response. We are now at an important crossroad whereby digital health, AI and genomic technologies are ripe for integration, clinical translation and implementation, and their application to infectious diseases represents an exemplar that could scale to many other areas of medicine.

Here, we apply our framework of automated data ingestion, preprocessing, quality control and standardisation^12,13^ in a real-world hospital context to input entire EHR data into multimodal fusion models for prediction of objective patient outcomes. As a proof of principle, we also developed a novel approach for microbial genome analysis and integration into a deep learning model using a genomic Large Language Model (gLLM) architecture and data standardisation using a proposed microbial genome Fast Healthcare Interoperability Resources (FHIR) format. This comprehensive fusion model of entire EHR and microbial genome data encompassed the SuperbugAI Platform and allowed us to evaluate the incremental benefit of the pathogen genome on clinical outcome prediction. Specific genomic features significantly associated with poor outcomes were then identified, providing important hypotheses for future biomarkers of disease, mechanistic insights into pathogenesis and identification of new therapeutic targets. This work provides novel approaches to the advances of multimodal AI systems for healthcare application, and the methodologies are scalable to all subspecialities of medicine where digital health and genomics will be used to personalise care.

## Results

### Patient population

We included a total of 2,656 hospitalisation episodes of confirmed bloodstream infection (BSI) involving 2,535 unique patients. The median age (range) of the patients was 65 (18 – 102) years, and the majority were male (61%) (Extended Data Table 1). There were 92 countries of birth, the majority (66%) being from Australia and New Zealand (Supplementary Figure S1). The most common comorbidities were diabetes mellitus (18.0%), followed by chronic renal disease (12.7%), heart failure (12.6%) and haematological malignancy (10.3%). The median (interquartile range) length of stay per BSI episode was 28 (8 – 51) days, and most patients (57.7%) had been previously hospitalised (last 12 months). A total of 8,380,179 structured EHR data points were used for the analysis, which included 12 months of EHR data preceding the onset of bloodstream infection. In addition, the unstructured clinical notes accounted for approximately 600 million distinct data tokens in total.

### Bloodstream Infections

The most common causative organism was the Gram-negative bacterial pathogen, *Escherichia coli* (n = 861, 58.2%) (Supplementary Table S1). Other Gram-negative bacteria included *Klebsiella pneumoniae* (n = 185, 13.5%), *Pseudomonas aeruginosa* (n = 145, 10.5%), *Proteus mirablis* (n = 52, 3.8%), *Acinetobacter species* (n = 39, 2.8%) and *Serratia marcescens* (n = 39, 2.8%). *Staphylococcus aureus* was the most common (n = 557, 44.5%) Gram-positive bacterial pathogen, followed by streptococcal species (n = 302, 25.3 %), *Enterococcus faecium* (n = 213, 17.9%) and *Enterococcus faecalis* (n = 139, 11.7%) (Supplementary Table S1).

### Clinical outcomes

In-hospital death from bloodstream infection (within 30 days of blood culture) was 9.4% (249/2,656), with 10.7% (276/2,572) patient episodes requiring ICU admission within 14 days of infection onset (Extended Data Table 2). Prolonged length of stay (≥ 14 days) was observed in 16.9% (448/2,656) of episodes and 6.8% (163/2,407) of discharges from hospital had an unplanned readmission within 30 days (Extended Data Table 2).

### Mortality

Using a multimodal EHR deep learning model, including all structured and unstructured data, the area under the receiver operating characteristic curve (AUROC) for predicting in-hospital death within 30 days of bloodstream infection was 0.93 (95% CI 0.924 - 0.942) (Figure 1). Performance was marginally higher for shorter horizons with AUROCs of 0.94 (95% CI 0.931–0.951) for 7-day and 0.94 (95% CI 0.926–0.952) for 14-day mortality. Extending the EHR data input window from one day post-infection to two- and three-days provided no improvement in performance (Supplementary Figure S2). These models significantly outperformed unimodal models that were trained exclusively on structured data alone from the EHR (AUROC ranging from 0.801 – 0.853), or a traditional approach for mortality prediction using the APACHE II scores (AUROCs 0.770 – 0.780) (Supplementary Figure S2).

**Figure 1.**
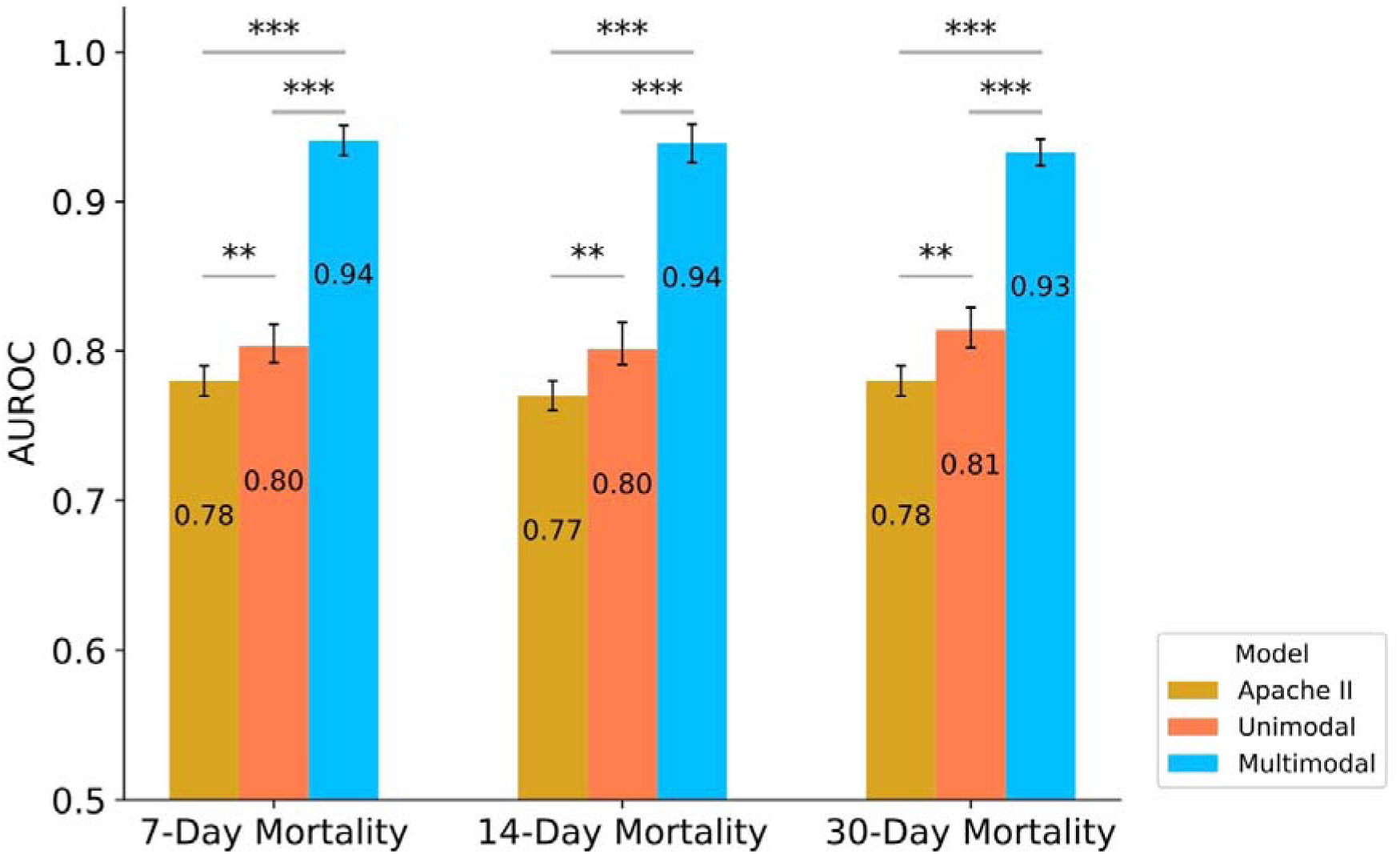
Mortality prediction of patients with bloodstream infection. Unimodal models were trained using structured electronic health record (EHR) data alone, whereas multimodal models were trained using complete EHR data (structured and unstructured) of the patient. All models were trained using data from the year preceding infection to 24hrs post positive blood culture draw. The performance of the models was compared against traditional mortality prediction with Apache II scores. Asterisks denote statistical significance (*: *P* value < 0.05; **: *P* value < 0.01; ***: *P* value < 0.001). The error bars represent 95% confidence intervals.

### ICU Admission

The multimodal deep learning model predicted ICU admission within 14 days of bloodstream infection with an AUROC of 0.978 (95% CI 0.966 - 0.986) (Table 1). Incorporating multiple data sources substantially improved predictive performance, with a clear increase in performance metrics compared to the unimodal (structured EHR only) baseline (AUROC 0.891 [95% CI 0.882 - 0.902]). The multimodal deep learning model performed similarly with inclusion of data one-, two- and three-days after a positive blood culture (data not shown).

**Table 1.**
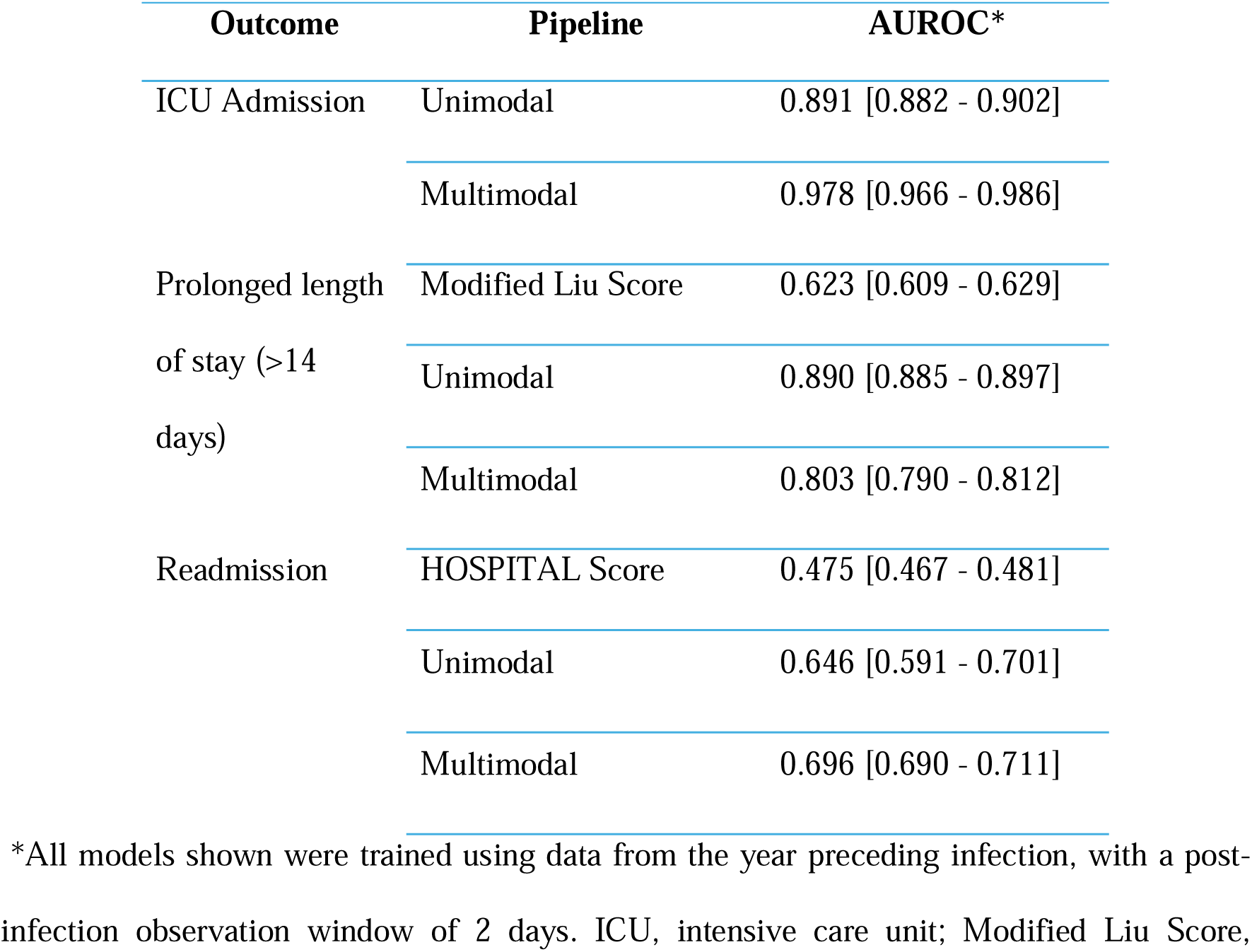
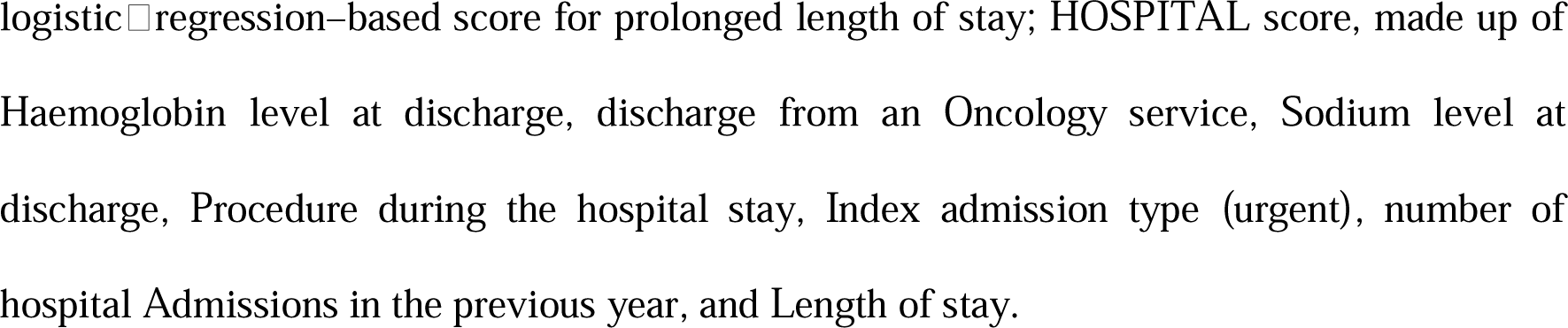
Model performance for clinical outcome prediction.

| Outcome | Pipeline | AUROC* |
| --- | --- | --- |
| ICU Admission | Unimodal | 0.891 [0.882 - 0.902] |
|  | Multimodal | 0.978 [0.966 - 0.986] |
| Prolonged length<br>of stay (>14<br>days) | Modified Liu Score | 0.623 [0.609 - 0.629] |
|  | Unimodal | 0.890 [0.885 - 0.897] |
|  | Multimodal | 0.803 [0.790 - 0.812] |
| Readmission | HOSPITAL Score | 0.475 [0.467 - 0.481] |
|  | Unimodal | 0.646 [0.591 - 0.701] |
|  | Multimodal | 0.696 [0.690 - 0.711] |

\*All models shown were trained using data from the year preceding infection, with a post-infection observation window of 2 days. ICU, intensive care unit; Modified Liu Score,
logistic regression–based score for prolonged length of stay; HOSPITAL score, made up of Haemoglobin level at discharge, discharge from an Oncology service, Sodium level at discharge, Procedure during the hospital stay, Index admission type (urgent), number of hospital Admissions in the previous year, and Length of stay.

### Length of stay

For predicting prolonged length of stay (≥ 14 days), the AUROC was 0.803 (95% CI 0.790 - 0.812) using our entire EHR multimodal deep learning model (Table 1). For this outcome measure, analysis using structured EHR data alone (unimodal) achieved higher prediction performance (AUROC 0.890 [95% CI 0.885 - 0.897]), indicating that LOS may be driven more by structured trajectory data (labs, vitals, LOS-relevant demographics) than by unstructured notes. An established length of stay prediction tool (modified Liu Score)^14^ was used as a comparator, and this achieved a significantly lower AUROC of 0.623 [95% CI 0.609 – 0.629] compared to both our AI approaches (Table 1).

### Unplanned Readmission

As an important hospital quality metric, we next predicted unplanned readmission within 30 days in patients who survived to discharge after an episode of bloodstream infection (n = 2,407). The multimodal deep learning model achieved an AUROC of 0.696 (95% CI 0.690– 0.711) for this outcome (Table 1). Unimodal modelling showed greater variability in performance, with an AUROC of 0.646 and wider confidence intervals (95% CI 0.591- 0.701). Prediction performance did not improve when extending the post-culture data window from one-day to two- or three-days. Nevertheless, our deep learning models significantly outperformed a traditional model for predicting unplanned readmission known as the HOSPITAL score^15^ (0.475 [0.467 - 0.481], Table 1).

### Digital health and pathogen genomics data fusion

To assess whether the genome of the bacteria causing the bloodstream infection in each patient could augment the predictive performance of our model, we developed the SuperbugAI platform for real-time integration of pathogen genome and EHR data for predictive patient-focussed analytics (Extended Data Figure 1). First, we proposed a FHIR based (version R5) standardisation of microbial genome data (Supplementary Figure S3) to allow for novel integration with entire EHR data. Next, we developed a gLLM approach for genomic representation in the model, which harnessed the semantic capture capabilities of natural language processing methodologies to capture biological context for modelling. This technique employed an approach to segment sequences into variable-length tokens (k-mers), effectively preserving the contextual information essential for interpreting the semantics of biological words. These tokens were then used as input data into the model.

As a proof of principle, we chose the two most common bacterial causes for bloodstream infection to evaluate our integrated prediction capacity; *E. coli* (n = 861) and *S. aureus* (n = 557). Extending our multimodal fusion approach, we combined unique patient EHR data with pathogen genomic gLLM tokens into the fusion model to predict 30-day mortality. The genomic datasets generated 247 million tokens for *E. coli* and 177 million tokens for *S. aureus*. Three model configurations were trained on structured EHR (Model 1), entire EHR (structured + unstructured) (Model 2), and entire EHR plus genomics (Model 3) (Supplementary Table S2). Compared to structured data alone (*E. coli* AUROC 0.7603 [95% CI 0.688 - 0.832], *S. aureus* AUROC 0.795 [95% CI 0.752 - 0.838]) or entire EHR data (*E. coli* AUROC 0.871 [95% CI 0.846 – 0.895], *S. aureus* AUROC 0.800 [95% CI 0.734 – 0.866]), the combination of entire EHR plus the pathogen genome data outperformed in the prediction of 30-day mortality for both *E. coli* (AUROC 0.917 [95% CI 0.879 - 0.955]) and *S. aureus* (AUROC 0.850 [95% CI 0.840 - 0.860]) bloodstream infection. Progressive improvement in performance (AUROC) was observed from Model 1 (structured EHR) to Model 3 (entire EHR plus genomics).

### Pathogen genome features influencing mortality prediction

To investigate the biological plausibility of our findings, we first examined the gLLM tokens that had the greatest influence on prediction performance based on attribution scores, which reflect how strongly each genomic token was considered in the prediction (attribution scores were derived from the model’s attention weights). To evaluate the discriminative value of the token attribution scores, tokens were grouped into high- (attribution score above the mean) and low-importance tokens (attribution score below the mean). A Chi-square test of independence revealed a statistically significant difference in mortality between patients infected with bacteria containing high versus low-importance tokens (*P* value < 0.001). As a secondary validation, Kaplan–Meier survival analysis^16^ further assessed the relevance of the token attribution scores with patient mortality, and showed that a high-risk signature, defined as the ten most highly ranked and discriminative tokens drawn from the total of 247 million and 177 million tokens in *E. coli* and *S. aureus*, respectively, showed a significant increase in mortality over time compared to patients infected with bacteria without the signature sequences (*P* value < 0.05; Supplementary Figure S4).

To uncover pathogenic insights and generate new hypotheses on bacterial virulence within human infection, the gLLM tokens were then mapped back onto the corresponding genomes. Aligning the gLLM tokens allowed a ranking of genes based on a weighting score that involved multiplying the proportion of independent tokens aligning per gene by the token’s attribution scores. Upon ranking genes based on their weighting score, we noted that virulence genes were strongly represented in the highest weighted genes whose scores were two standard deviations above the mean value for both *E. coli* (92 genes) and *S. aureus* (57 genes) (Supplementary Table S3 and Supplementary Table S4, respectively). Gene enrichment analysis of the highest weighted genes showed that for *E. coli*, enrichment in iron acquisition pathways related to transmembrane transport and import of iron was observed. These pathways are crucial for the survival and virulence of pathogenic bacteria.^17,18^ Representative genes were identified from the *fhu* operon (*fhuB*, *fhuC*, and *fhuE*) (Figure 2A), which encodes an ABC transporter system (Figure 2B)^19,20^ that plays a critical role in acquiring iron containing siderophores within host environments,^20,21^ and the *fep* operon (*fepA*, *fepC* and *fepD*), which is involved in the uptake of ferric-enterobactin, a high-affinity siderophore important for iron acquisition and scavenging under low iron conditions such as during infection.^22^ Of these genes relating to iron acquisition, *fhuB*, which encodes an integral membrane channel protein (Figure 2B), had one of the highest weighted scores (Supplementary Table S3).

**Figure 2.**
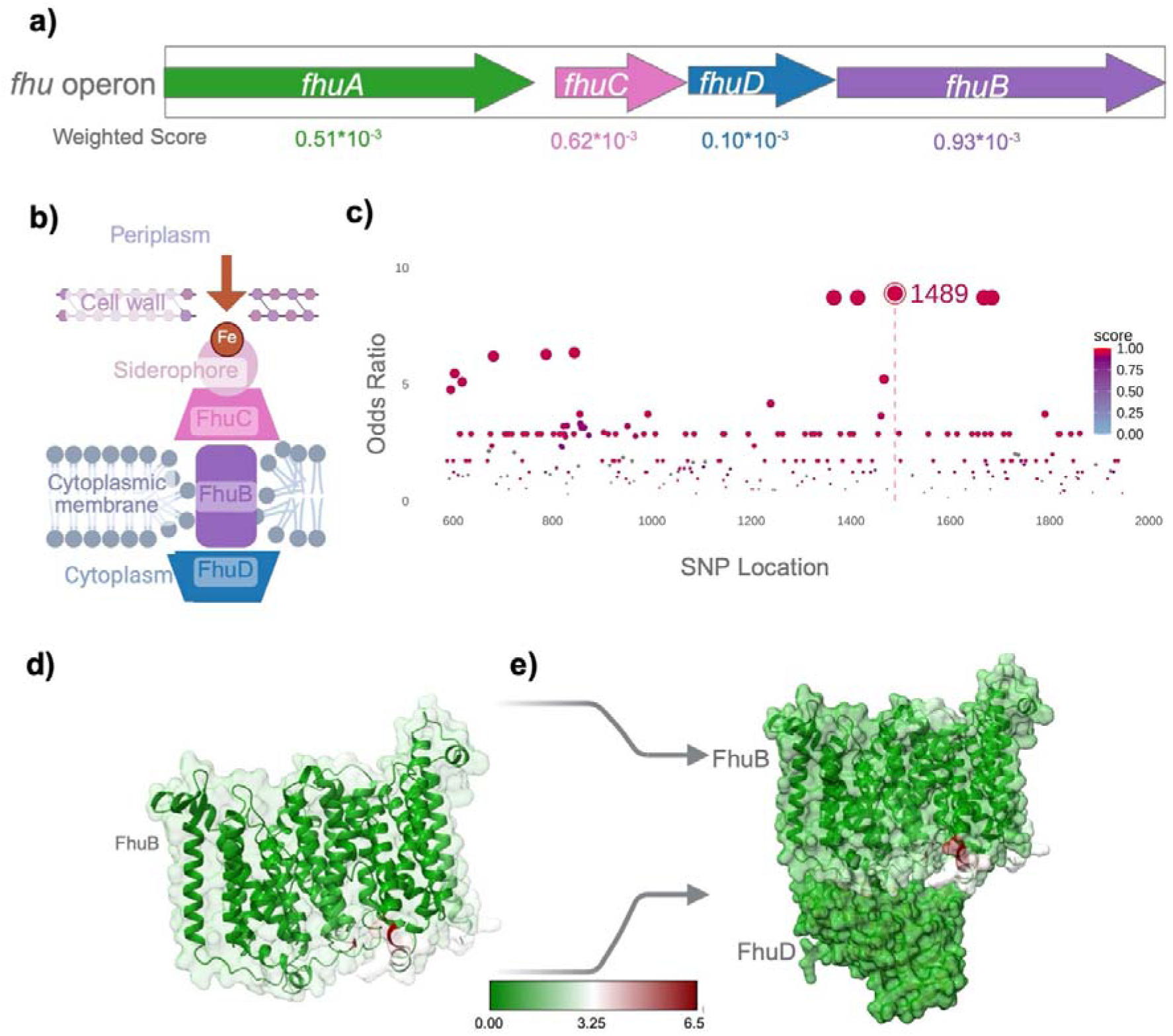
*fhu* operon influencing patient mortality from *E. coli* bloodstream infection. [A] Schematic representation of the *fhu* operon, showing the four constituent genes *fhuA-B*. Arrows indicate the direction of transcription. The weighted score for each gene is shown below the corresponding arrow. Higher scores indicated greater contribution to the 30-day mortality. [B] Schematic diagram showing the organisation of the FhuD (periplasmic substrate-binding protein), FhuB (inner membrane permease), and the FhuC (cytoplasmic ATP-binding energy-providing component) ferric iron (Fe³) transport system. [C] Plot showing odds ratios for the SNPs within *fhuB* associated with 30-day mortality, as identified by treeWAS analysis. The highlighted SNP at position 1489 had the highest odds ratio for patient mortality. [D] Predicted structure of the FhuB protein generated in-silico using AlphaFold 3. The structure is color-coded according to the per-residue Ca RMSD (Root Mean Square deviation between the alpha-carbon atoms) profile, illustrating structural deviations between the wild-type and the mutated proteins. [E] Predicted secondary structure of the FhuB–FhuD protein complex showing that the structural deviations within FhuB are localized to regions involved in interactions with FhuD. (Image created using BioRender.com)

As *fhuB* is part of the *E. coli* core genome and present in all the infecting isolates, we aimed to identify specific markers within the *fhuB* sequence that were associated with mortality. We aligned all *fhuB* sequences from patients who died and those who survived and performed a sequence comparison and SNP analysis (Figure 2C). Notably, we identified a SNP resulting in an amino acid (aa) substitution (T1489C) that was significantly associated with 30-day patient mortality. Studies have shown that point mutations within *fhuB* can have function-modulating effects in *E. coli*,^23^ as well as in other bacterial species.^24^ Comparing the predicted protein structure for the mortality-associated aa substitution and the wild-type FhuB^25^, we identified significant FhuB transmembrane helical structure alterations in isolates associated with mortality (Figure 2D). These structural changes were in areas where FhuB specifically interacts with FhuD, which provides the energy to drive transport of iron- siderophore complexes into the cell (Figure 2E).^26^ We hypothesise that these changes in FhuB may affect iron uptake of the bacteria and may account for the strong association of this protein to the mortality of patients with *E. coli* bloodstream infection.

For *S. aureus*, we similarly identified virulence related genes as top-ranking based on weighting scores, and this included a group of genes associated with the Ess system, including *esxB*, *essB* and *essC* (Figure 3A). This system, also referred to as the Type VII secretion system (Figure 3B), is also part of the *S. aureus* core genome and is implicated in staphylococcal pathogenesis by inferring bacterial competitiveness and modulating the host immune response.^27^ We identified that *essC* was the highest ranked gene based on weighted score, and as performed for *E. coli*, we compared all the *essC* sequences from patients who survived with those who died to identify significant SNPs that were highly associated with 30-day mortality (Figure 3C). We identified a large co-occurring SNP cluster (Figure 3C inset), and its secondary structure was compared to the wild-type variant. The predicted protein structure (Figure 3D) revealed a notable change in the EssC protein (RMSD of 6.883 Angstrom), most pronounced within the transmembrane helical structures and the Cytoplasmic 2 domain, which includes two forkhead-associated structures important for signal transduction and transcriptional regulation.^28^ In contrast, the Cytoplasmic 1 domain, critical for substrate specificity in Type VII secretion,^29,30^ remained relatively unchanged (Figure 3D).

**Figure 3.**
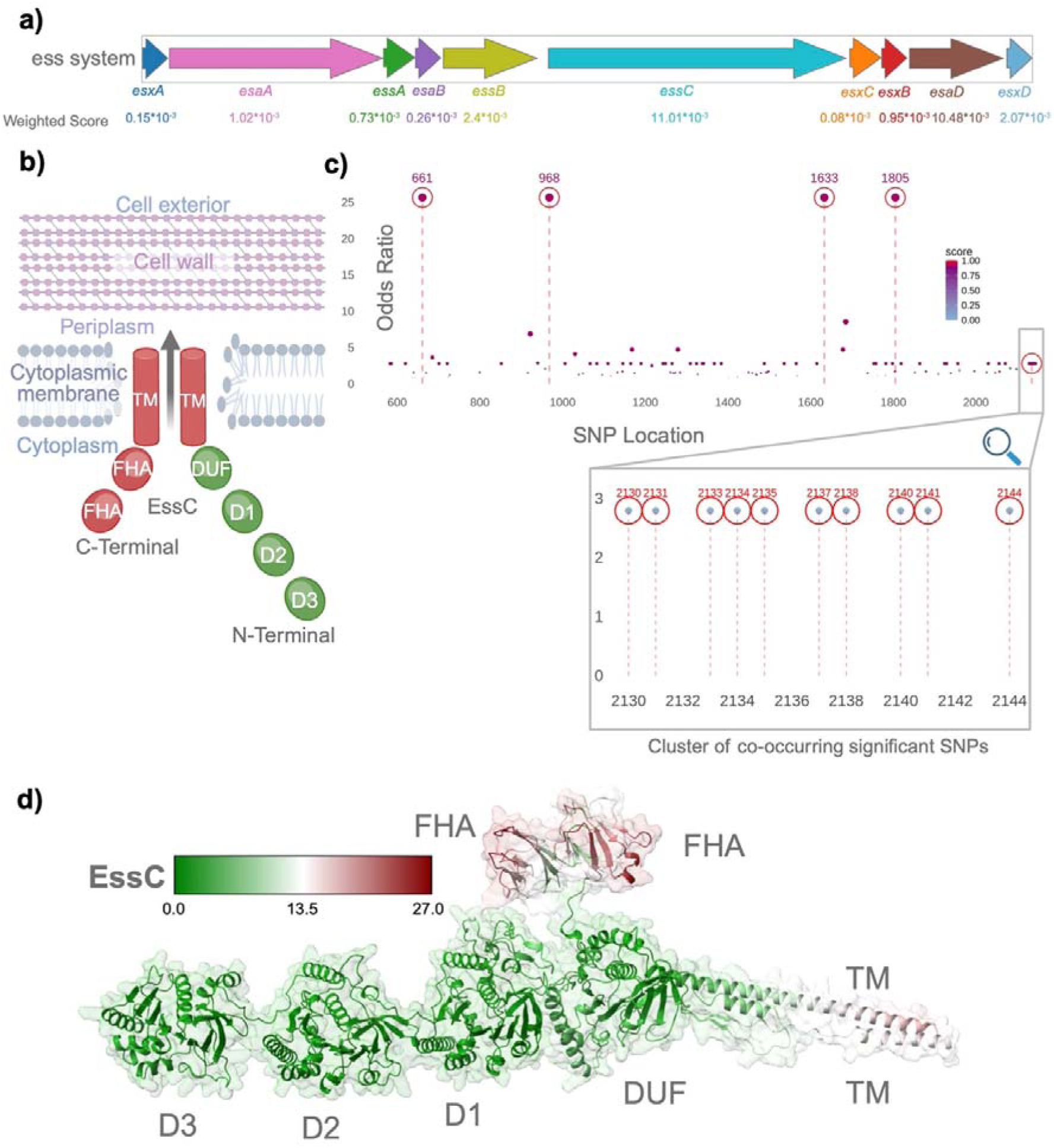
Ess system influencing patient mortality from *S. aureus* bloodstream infection. [A] Schematic representation of the *ess* operon showing the constituent genes. Arrows indicate the direction of transcription. The weighted score for each gene is shown below the corresponding arrow. Higher scores indicate greater contribution to 30-day mortality. [B] Schematic showing the different functional domains of the Ess protein and their organisation. The N-terminal region comprises the ATPase domains D1, D2, and D3 and the C-terminal contains forkhead-associated (FHA) domains. The transmembrane (TM) helices are also depicted, which constitute a key structural component of the T7SS machinery. [C] Plot showing attribution scores and odds ratios for the SNPs within the *essC* gene associated with 30-day mortality, as identified by treeWAS analysis. [D] Predicted secondary structure of the EssC protein generated using AlphaFold 3. The structure is color-coded according to the per-residue Root Mean Square deviation between the alpha-carbon atoms (Ca RMSD) profile, illustrating structural deviations in the FHA and TM domains between the wild-type and the mortality-associated mutated protein. (Image created using BioRender.com)

Taken together, these data suggest that our gLLM tokens associated with patient mortality from *E. coli* and *S. aureus* bloodstream infection could be linked to specific virulence-related proteins and predicted structural changes that could provide biological plausibility for their role in predicting patient mortality in the context of entire EHR data.

## Discussion

Multimodal AI applications have the potential to transform the future of healthcare and leverage the highly diverse data that is now encountered in patient care. Deep learning models that not only have the capacity to incorporate the complexity of the entire EHR but are also able to ingest ancillary health data such as genomics, represents the future of AI in healthcare. Here, we have applied an automated data ingestion pipeline for quality control and pre-processing of the entire EHR, involving over 608 million pieces of data, to accurately predict objective patient outcomes from life-threatening infection. Most notably, we also developed an exciting methodological approach using a gLLM to incorporate pathogen genome data from the causative bacteria with the entire EHR data from a given patient in multimodal AI fusion models and showed augmented prediction performance for patient mortality. Interrogation of the most predictive parts of the bacterial genome led to biologically plausible mechanisms for human disease and highlights the power of this approach to uncover and prioritise pathogenic mechanisms that are most relevant to human infection. The described multimodal AI fusion approaches are also scalable to any area of human health whereby entire patient data and genomics (human or pathogen) are relevant for disease prediction, diagnosis, risk stratification and precision treatment.

From 2,656 hospitalisation episodes of confirmed bloodstream infection, inclusion of both structured and unstructured EHR data outperformed unimodal structured data alone and traditional models for predicting objective patient outcomes. This was most pronounced for predicting mortality and the need for ICU admission. Inclusion of unstructured data remains challenging globally as healthcare notes may not always be included in electronic health systems, and when they are, the data is often stored differently in data warehouses, conversion to a data standard is more complex and therefore impacts interoperability, and the handling of such data within a secure environment is often essential due to protected health information. Here, we applied our newly developed digital health toolbox that allows for complete automation of unstructured (and structured) data pre-processing and conversion to OMOP or FHIR, as well as quality control and handling of data anomalies prior to input into deep learning models. This data ingestion and pre-processing pipeline is essential for healthcare to fully leverage the power of EHR data and AI applications, is agnostic to EHR service provider and has the potential of being scalable to all disciplines of medicine.

Like AI, genomics is another technology that is transforming healthcare. The application of human genomics to precision care for oncology, neurology, pharmacology and genetic diseases, is increasingly established.^31,32^ More recently, AI applications using human genome data to support patient care have been reported,^2,32,33^ and HL7 FHIR standards for human genome data are now defined.^34,35^ For infectious diseases, pathogen genomics is emerging as an important tool, but has mainly been used for tracking outbreaks, identifying transmission events and detecting antimicrobial resistance genes.^36–38^ Applications for direct patient care outside of antimicrobial resistance has been limited,^39^ despite the wealth of research that has gone into studying microbial pathogenesis within animal infection model systems.^40,41^ Here, we present the first report of an HL7 FHIR standard for microbial genome data, and the development of a new gLLM approach to input these data into multimodal deep learning fusion models. We showed that for two of the most common causes of bloodstream infection, the addition of the pathogen genome to the entire EHR data of a given patient improved the performance of our AI models in predicting patient mortality. This hypothesis-agnostic approach of genome data input using a gLLM was then interrogated, and most notably, identified specific bacterial mechanisms that had strong plausibility for impacting human disease. The power of this approach is that it provides unique insights into bacterial virulence within a human, tightly linking the full genetic code of the specific infecting bacteria with the features of the human host as represented by the specific EHR data of a given patient. These insights could have important implications for the study of bacterial pathogenesis, identification of novel therapeutic targets and diagnostics to support risk stratification and bacterial virulence assessment. Furthermore, the approaches used for integrating these diverse data types are scalable to other infectious diseases, and other areas of medicine where genomics, including human genomics, and clinical data integration could help support precision medicine. These proof of principle results strongly support further research using larger patient numbers, patients from different institutions and different infecting pathogens, as well as other areas of medicine that utilise genomics for patient care.

We are now at an exciting crossroad whereby AI, genomics and digital health are ripe to transform the future of healthcare. To fully realise the potential of these transformative technologies, EHR systems and AI Health applications must evolve to manage the complexity of genomic information while addressing key challenges such as patient autonomy, access, genetic literacy, privacy, and data transferability. These approaches have real potential to harness the full magnitude and complexity of high dimensional clinical data for truly individualised care.

## Methods

### Patient population and cohort selection

All consecutive patients with bloodstream infection from The Alfred Hospital, Melbourne, Australia between 2018 – 2023 were included in the study. Patients with coagulase-negative staphylococci in their blood were excluded due to their unclear clinical significance. The time of onset of the bloodstream infection was defined as the date and time of the first positive blood culture draw (time 0). All included patients had the causative bacterial pathogen stored within a prospective biobank −80°C. This study was approved by the Alfred Hospital Ethics Committee (Project No. 185/21).

### Microbiology and Pathogen Genomics

All included isolates were identified and underwent antimicrobial susceptibility testing within the hospital clinical microbiology laboratory using Matrix-Assisted Laser Desorption/Ionization Time-of-Flight (MALDI-TOF) and VITEK2, respectively. Genomic DNA was extracted from each isolate using the GenFind v3 kit, and whole-genome sequencing (WGS) was performed on the Illumina NovaSeq platform. Genomes were assembled with Unicycler v0.5.0^42^ and annotated using Bakta v1.8.1 with database v5.0.^43^ We identified species for each genome with PathogenWatch v3.0.1.^44^ Genomes with <20x depth, with assembly size >20% more than the expected genome size for their species, or genomes where the assigned genus did not match the MALDI-TOF identification,^45^ were excluded from further analysis. All *E. coli* and *S. aureus* genomes used in this study have been uploaded to the public archives (bioproject number: PRJNA1493863).

### EHR Data and pre-processing

Raw EHR data for each admitted episode of bloodstream infection was extracted from the hospital data warehouse. During data extraction, the EHR records were de-identified to safeguard patient confidentiality. Structured EHR data included static demographic information and temporal laboratory and vital sign measurements. Unstructured EHR data included all the healthcare notes in English text format. We adopted two international open data standards to represent the EHR data; OMOP-CDM^46,47^ and FHIR (Fast Healthcare Interoperable Resources).^48^

### Structured EHR data

The EHR data was standardised using the EHR-QC^12^ standardisation module. This process involved formatting the data to OMOP-CDM and mapping clinical concept names to Systematized Nomenclature of Medicine (SNOMED)^49^ and International Classification of Diseases (ICD-10)^50^ terminologies. The EHR data was extensively explored, underwent quality assurance and automated handling of data anomalies such as outliers, missing data, and multiple representations of the same data type, and was pre-processed using our in-house quality assurance module of the EHR-QC pipeline^12^ (Supplementary Methods S1; and Supplementary Figure S5).

### Unstructured data

Natural language processing (NLP) methods^51^ were adapted to get insight into the clinical notes. For the clinical notes pre-processing, we curated document categories including admission, progress, consulting, radiology and pathology notes by chronologically concatenating them at the admission episode level. Our analysis involved constructing a sub- word model^52^ from scratch using PubMed and our site-specific clinical notes. Pubmed data was retrieved, and abstracts and article titles were processed. To construct an Alfred hospital- specific word embedding model for the blood stream infection cohort, document titles of clinical notes underwent manual inspection and selection, discarding any irrelevant document categories. MedscispaCy^53^ was used for domain specific tokenisation of notes. We generated token-level embeddings following the BioWordVec methodology^52^ and separately generated document-level embeddings for each note using Doc2Vec^54^. Both models were then fine-tuned for clinical outcome predictions on the bacteraemia cohort. In our exploratory benchmarking, the token embeddings were found to be more informative and were used for downstream analyses.

### Clinical Outcome Measures

As proof of principle, we chose four objective clinical outcome measures to evaluate the predictive capacity of the SuperbugAI models. For all outcome measures, time zero was defined as the time the positive blood culture was drawn. Mortality was defined as all-cause in-hospital mortality and we evaluated predictions for 7, 14 and 30 days post time-zero. Admission to ICU was defined as the need for intensive care during the first 14 days from the positive blood culture draw (time zero). An ICU admission was considered true if a patient was first admitted to a general ward and then transferred to the ICU, with the transfer occurring after the blood culture draw (time zero) but within 14 days. Patients already in ICU at time-zero were excluded from the ICU-admission outcome cohort. Prolonged length of stay was defined as hospital stays longer than 14 days after the positive blood culture draw (time zero). Unplanned readmission was defined as readmission within 30 days following discharge that was not scheduled at the time of discharge. For all predictive analytics of clinical outcomes, an EHR data input window of 365 days prior and one-, two- and three-days post bloodstream infection (date of blood culture draw) was used.

### Deep learning models

We employed two distinct modelling approaches to predict clinical outcomes. The first approach utilised a multimodal architecture that integrated four different types of data inputs (Figure 4). For baseline comparison, we used a published unimodal architecture that included only structured data and composed of an ensemble of diverse individual machine learning algorithms^13^.

**Figure 4.**
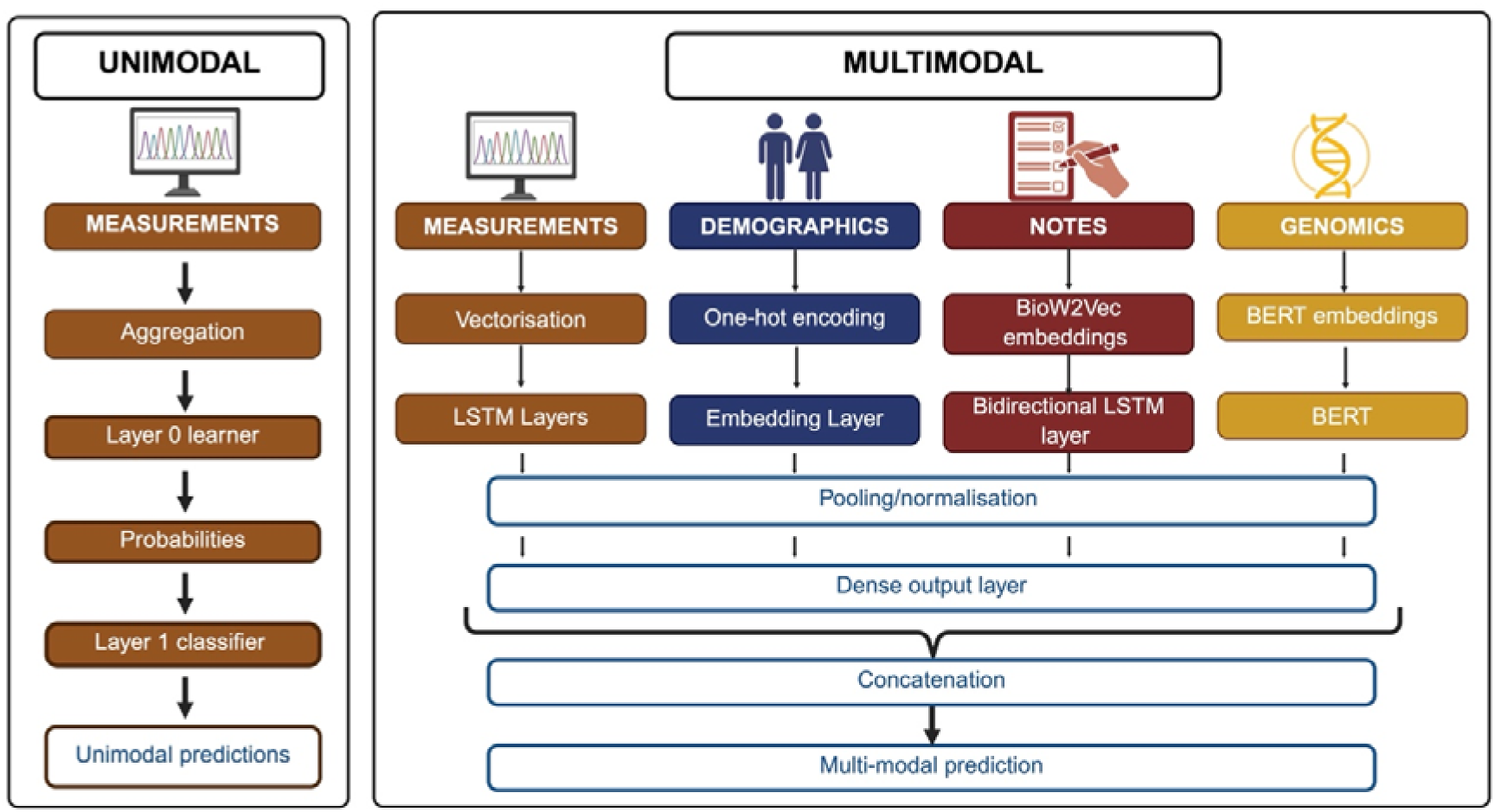
Deep Learning Architecture. The unimodal ensemble architecture combines multiple machine learning models at the first level, referred to as Layer 0 learners, whose outputs are then combined by a Layer 1 classifier to generate the final prediction. Multi- modal fusion includes diverse data types, such as measurements, demographics, clinical notes, and genomic data, which were processed through individual neural network stages, followed by late-stage fusion to produce the final prediction output. The preprocessing stages included aggregation, vectorisation, one-hot encoding, embedding techniques (BioW2Vec and BERT) and normalisation. A fusion of modalities was performed at the concatenation layer, ultimately feeding into a dense output layer for prediction. (Image created using BioRender)

### Uni-modal ensemble model

The unimodal model included the structured temporal data as a single modality, consisting of vital signs and laboratory measurements obtained from the EHR. We earlier designed an ensemble learner to specifically handle the intricacies of EHR data^13^. The ensemble was structured into two layers such that the probability outputs from the first layer formed the input to the second layer to produce the outcome. We included four distinct machine learning classifiers for each feature group (i.e., vitals, measurements and static) in the first layer and one in the second layer to combine all the intermediate outputs. During the training process, the hyper-parameters of all the individual learners were tuned individually. A hierarchical aggregation of feature importance scores was performed, where a weighted sum of the feature importance scores from the two layers were obtained to help interpret the ensemble architecture. The data was partitioned chronologically by admission time into 85:15 training and test sets, respectively.

### Multi-modal fusion

For the multimodal deep learning model, we built EHR-fusion, a late fusion model using the long short-term memory (LSTM)^55^ neural network architecture to combine data from three or four modalities (Figure 4). In the three-modality mode, we used static EHR (demographics) data, temporal EHR measurements and the unstructured clinical notes as input. For the evaluation of the incremental benefit of adding microbial genomics data to prediction performance, we overlayed the genomic information as the fourth modality input. The LSTM architecture was selected based on data sizes and for its efficiency in processing time series data. The static EHR data was processed using one-hot encoding combined with an embedding layer, while the numerical vectors representing measurements were processed using a unidirectional LSTM. Clinical notes were represented using sub-word embeddings and processed with a bidirectional LSTM. Genomic tokens were generated using the gLLM. We opted for a late fusion approach where different modalities were encoded and processed separately using neural networks, which was then fused using a concatenation layer and fed into a binary classification layer (Figure 4). The data was organised chronologically by admission time and partitioned such that 85% of the data, representing earlier patient episodes, formed the training set, while the remaining 15%, which included the most recent episodes, constituted the testing set. This chronological split allowed for the model to always be trained on past data and evaluated on future (unseen) data. Crucially, to prevent data leakage, we kept all episodes from the same patient within the training set if their initial episode fell into that partition.

### Model evaluations

Within the training partition, model hyperparameters were tuned using 5-fold cross-validation; final performance metrics (balanced accuracy, F1 score, AUROC) were then computed on the held-out chronological test set, with mean, standard deviation and 95% CI reported across the 5 folds’ validation performance for tuning stability, and a single point estimate with bootstrap 95% CI reported for the held-out test set. In benchmarking analysis, the performance of unimodal models in predicting the outcome measures was compared to traditional prediction systems, such as the APACHE II^56^ score for mortality, the modified Liu Score^14^ for prolonged length of stay, and the HOSPITAL Score^15^ for unplanned readmission. No established comparator was found for the need for ICU outcome, so this outcome was excluded from benchmarking analysis. Benchmarking comparisons used AUROC with 95% confidence intervals as the primary metric.

### Genomic Large Language Model (gLLM)

Assembled genomic contigs for the *E. coli* and *S. aureus* genomes were initially broken down into smaller units called tokens (Figure 5), analogous to biological words. While inspired by traditional k-mers, our approach involved a unique path by building a gLLM. Genome tokenisation was performed using the Unigram tokenizer (Eqn1-2), a data-driven approach that treats tokens as independent units and optimises vocabulary selection by computing token probabilities across the corpus and minimising a defined loss function.

**Figure 5.**
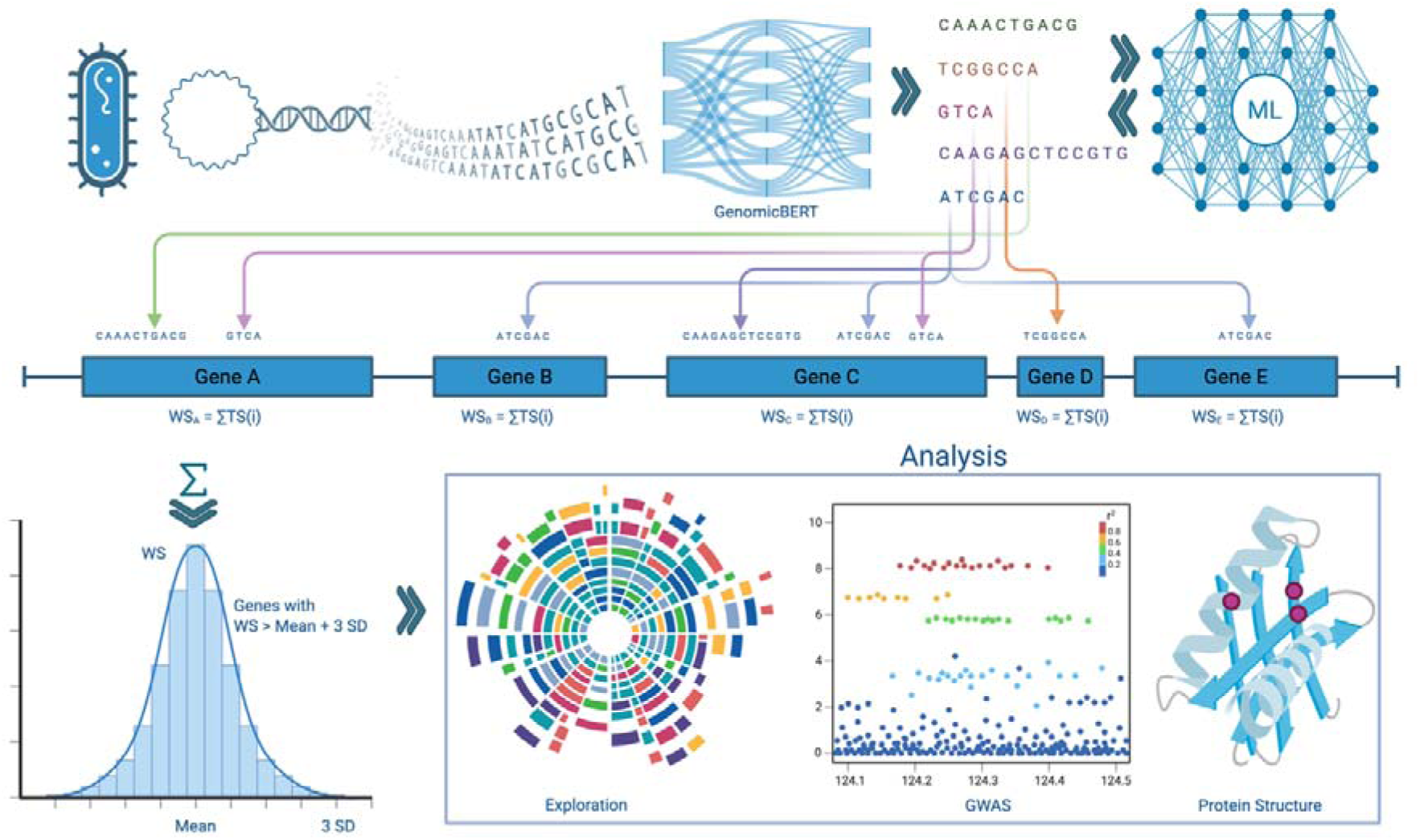
Schematic representation summarising the deep learning-based genomic data analysis workflow for the SuperbugAI Platform. The schematic outlines key steps: bacterial genome extraction, sequence tokenisation into k-mers, predictive modelling, genes of high importance calculation, and downstream evaluation including pangenome exploration, GWAS analysis, and secondary structure generation and comparison. (Image created using BioRender.com)

The probability of a token or sub-word sequence *x* = (*x*_1_*,…,x_M_*) is defined as:

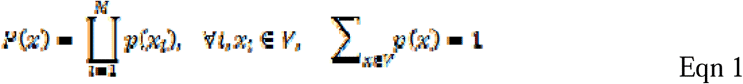

via Expectation-Maximization by maximising

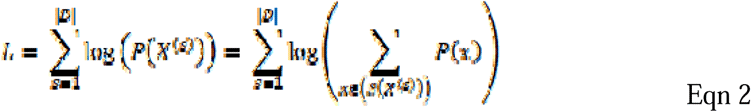

We then used these tokens to construct a genome-specific language model comprising 22 encoder layers, each with a hidden size of 768, 12 self-attention heads, and an intermediate feedforward network size of 1,152. The gLLM was pretrained using a masked language modelling and causal next-token prediction objective. The model contained 113,403,648 trainable parameters in total. The models were optimised for efficiency and scalability. It incorporates five architectural innovations: Rotary Positional Embeddings (RoPE), which encode positional information directly within the attention mechanism; GeGLU activation, which introduces a gating mechanism to improve feature learning and training stability; alternating local-global attention; unpadding; and FlashAttention3. Together these enabled effective modelling of long-range genomic dependencies while reducing computational cost and memory overhead during training and inference.

To study the biological significance of gLLM tokens (Figure 5), we mapped them back onto their respective genomes based on their positional coordinates. Each token was assigned an attribution score (using attention-weight aggregation) with respect to the mortality prediction output of the fine-tuned model. High-scoring tokens were defined as three standard deviations above the mean. A weighted score (WS) was also calculated for each annotated gene, which was the sum of each token’s attribution score multiplied by its frequency of occurrence and normalised by gene length (number of tokens overlapping the gene). High-scoring genes were defined as having an aggregated weighted score of three standard deviations above the mean. Kaplan–Meier survival analyses^16^ with the log rank test were conducted for survival at 30-days, comparing patients infected with bacteria that carried high scoring genes with the others.

The functional relevance of high scoring genes was evaluated by cross-referencing with curated virulence and antimicrobial resistance databases, namely AMR FinderPlus^57^, VFDB^58^, and the VF (Virulence Factor) collection^41,59^. For the highest scoring genes for *E. coli* and *S. aureus*, a genome-wide association study (GWAS)^60^ was performed using treeWAS^25,61^ to identify SNPs in these genes significantly correlated with 30-day in-hospital mortality. The predicted impact of the SNPs on the encoded protein was then evaluated using AlphaFold 3^25^, comparing wild-type proteins with the variant proteins and plotting the structural variations at the individual amino acid level. The overall protein variation was quantified by the average RMSD^62^ and the structural domains involved were predicted to infer the potential underlying pathogenic mechanisms associated with mortality.

### Integrated analytic platform set up

The structured EHR data was hosted on a virtual machine (VM) within the Microsoft Azure secure cloud, referred to as the Central VM (Extended Data Figure 1). A PostgreSQL server was deployed to store the standardised EHR records in a relational database. Simultaneously, the data was also loaded onto the FHIR server maintaining synchronisation between the two systems. Using both FHIR and OMOP standards ensured that the EHR data was both interoperable (FHIR) and well formatted for analytical purposes (OMOP), enabling seamless real-time analytics. A seamless synchronisation was achieved between these two data stores using bi-directional mapping. This information was harmonised with pathogen genomic data to provide a robust platform capable of integrated representation and analyses (Supplementary Methods S2). The platform worked seamlessly with in-house tools enabling fully automated data quality-control and processing. Further, it facilitated live query and linked data export, enabling harmonisation and data modelling. It also included interactive dashboards for exploration and analysis. The entire infrastructure (Extended Data Figure 1) was secured behind a firewall, with external access restricted to the interactive dashboard, which was accessible only within the hospital network.

## Data Availability

All data produced in the present study are available upon reasonable request to the authors

## Acknowledgements

The authors acknowledge funding support of Medical Research Future Fund (MRFF) for the SuperbugAI flagship project (Grant FSPGN000048). YR received the Monash Graduate Scholarship for his PhD. We thank Tyrone Chen and Jasbir Dhaliwal for their contributions in the initial phase of the project, and Naima Vahab for her support with the gLLM. This work was supported by Monash eResearch capabilities, including the high-performance computer M3 and Research Data Storage. We acknowledge the use of graphical icons sourced from Flaticon (flaticon.com) in the figures.

## Declarations

Nil.

## Extended Data

**Extended Data Figure 1:**
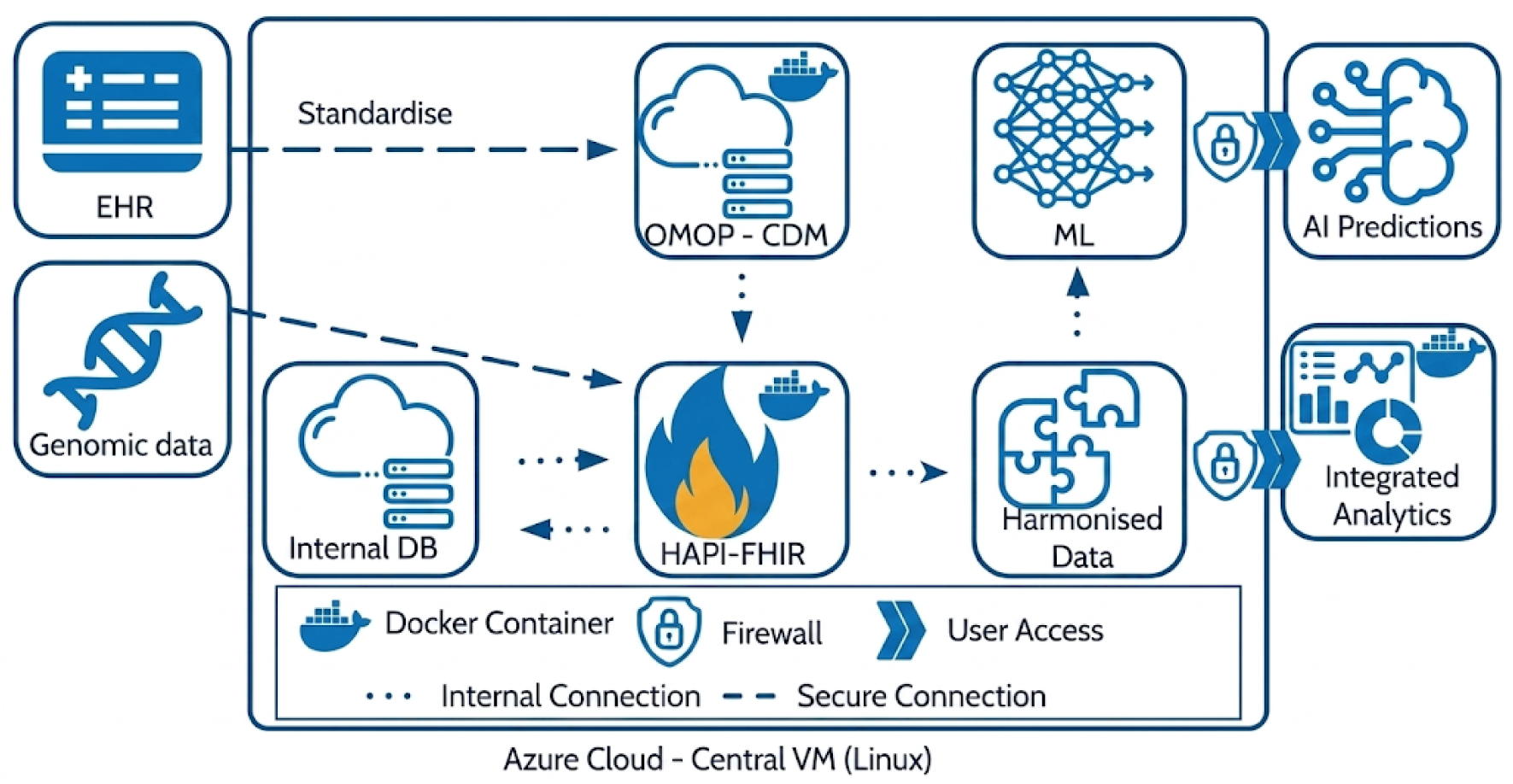
SuperbugAI data storage and analytics platform. Input data included the entire institutional EHR and the genome sequence data of the cultured causative bacterial pathogens. The infrastructure included a central Azure cloud VM, a PostgreSQL database, HAPI-FHIR server along with its internal data-store, and the harmonised data. The harmonised data provided the integrated representation of EHR, and genomic information used for analytical and modelling purposes. The entire infrastructure was secured within the hospital firewall.

**Extended Data Table 1.**
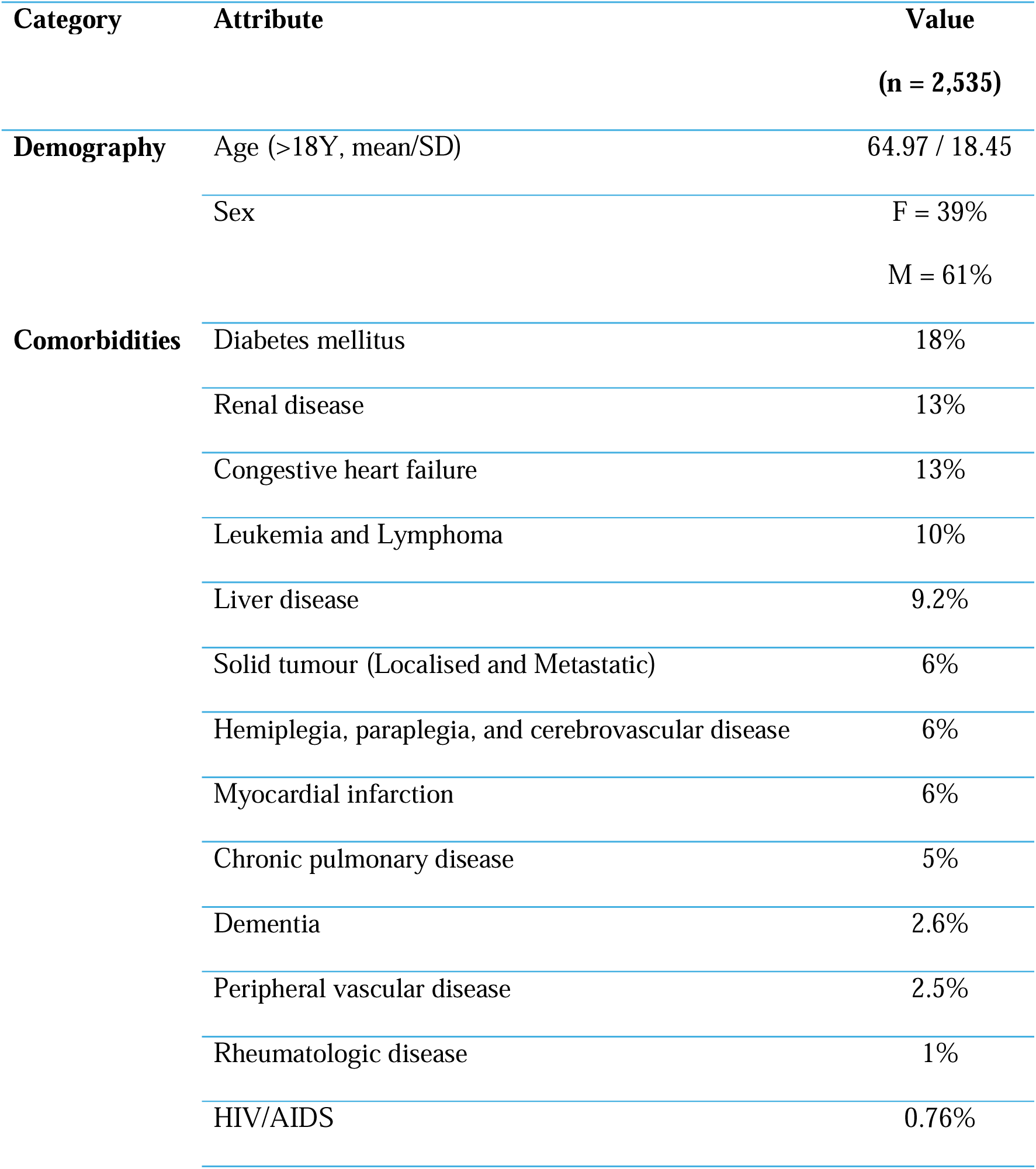
Patient Characteristics.

| Category | Attribute | Value |
| --- | --- | --- |
|  |  | (n = 2,535) |
| Demography | Age (>18Y, mean/SD) | 64.97 / 18.45 |
|  | Sex | F = 39% |
|  |  | M = 61% |
| Comorbidities | Diabetes mellitus | 18% |
|  | Renal disease | 13% |
|  | Congestive heart failure | 13% |
|  | Leukemia and Lymphoma | 10% |
|  | Liver disease | 9.2% |
|  | Solid tumour (Localised and Metastatic) | 6% |
|  | Hemiplegia, paraplegia, and cerebrovascular disease | 6% |
|  | Myocardial infarction | 6% |
|  | Chronic pulmonary disease | 5% |
|  | Dementia | 2.6% |
|  | Peripheral vascular disease | 2.5% |
|  | Rheumatologic disease | 1% |
|  | HIV/AIDS | 0.76% |

**Extended Data Table 2.** Summary of clinical outcomes.

| Clinical Outcome | Proportion of BSI Episodes |
| --- | --- |
|  | (Count/Total %) |
| Mortality within 30 days of BSI | (249/2,656) 9.37% |
| Prolonged LOS (≥ 14 days) | (448/2,656) 16.89% |
| Need for ICU within 14 days from BSI | (276/2,572) 10.73% |
| Unplanned readmission within 30 days of discharge | (163/2,407) 6.82% |
| BSI; Bloodstream infection, ICU; Intensive care unit, LOS; Length of stay. |  |

